# Indoor residual spraying practises against *Triatoma infestans* in the Bolivian Chaco: contributing factors to suboptimal insecticide delivery to treated households

**DOI:** 10.1101/2020.12.21.20248629

**Authors:** Raquel Gonçalves, Rhiannon A.E. Logan, Hanafy M. Ismail, Mark J.I. Paine, Caryn Bern, Orin Courtenay

## Abstract

**Background:** Indoor Residual Spraying (IRS) of insecticides is a key method to reduce transmission by *Triatoma infestans*, vector of Chagas disease in a large part of South America. However, the successes of IRS in the Gran Chaco region straddling Bolivia, Argentina and Paraguay, have not equalled those in other Southern Cone countries.

**Aims:** This study evaluated routine IRS practises and insecticide quality control in a typical endemic community in the Bolivian Chaco.

**Methods:** Alpha-cypermethrin active ingredient (a.i.) concentrations captured onto filter papers fitted to sprayed wall surfaces, and in prepared spray tank solutions, were measured using an adapted Insecticide Quantification Kit (IQK™). The results were analysed by negative binomial GLM regression in relation to the time (minutes) spray teams spent treating houses, spray rates (surface area to spray [m^2^/minute]), householder compliance to empty houses for IRS delivery, and the visual presence/absence of filter papers. The IQK™ assays developed for these samples were validated against HPLC quantification methods. Results: Substantial variations in the delivered a.i. concentrations were observed; only 10.4% (50/480) of filter papers, and 8.8% (5/57) of houses received the target concentration. The delivered concentrations were not related to those in the matched spray tank solutions. The sedimentation of a.i. in the surface solution of prepared spray tanks was rapid, resulting in a 29% loss of a.i. content within 5 minutes, and 48.5% after 15 minutes. The delivered concentrations were positively associated with the time spent spraying the house, and inversely related to the spray rate, but showed weak correlations in both cases. The influence of householder compliance on spray rates were significant, though associated differences in delivered concentrations were not detected. No differences were observed in spray rates between houses fitted with filter papers and houses without.

**Conclusions:** Suboptimal delivery of IRS is partially attributed to the insecticide physical characteristics, and the need for revision of insecticide delivery methods, which includes training of IRS teams and community education to encourage compliance. The IQK™ is a necessary field-friendly tool to improve IRS quality, and to facilitate health worker training and decision making by the Chagas disease vector control managers.

## Introduction

Chagas disease results from infection with the parasite *Trypanosoma cruzi* (Kinetoplastida: Trypanosomatidae) which causes a range of pathologies in humans and other animals. In humans, acute symptomatic infection occurs within weeks to months of infection, characterised by fever, malaise, and hepatosplenomegaly. An estimated 20-30% of infections progress to chronic forms, most commonly cardiomyopathy, characterized by conduction system deficits, arrhythmias, left ventricular dysfunction and eventually congestive heart failure, and less frequently by gastrointestinal forms of the disease. These conditions develop over decades, and are difficult to treat [1]. There is no vaccine.

The estimated global burden of Chagas disease in 2017 was 6.2 million resulting in 7.9 thousand deaths, and causing 232 thousand all-age Disability-Adjusted Life-Years (DALYs) [2, 3, 4]. *T. cruzi* is transmitted by triatomine bugs (Hemiptera: Reduviiidae) throughout Central and South America, and parts of southern North America, which accounted for 30 thousand (77%) of the total new cases in 2010 in Latin America [5]. Congenital transmission and infected blood transfusions are additional routes of infection, occurring in non-endemic regions such as Europe and USA. In Spain, for example, there are an estimated 67,500 infections among Latin American immigrants[6], at an annual cost to the healthcare system of % 9.3 million USD [7]. In a Barcelona hospital, between 2004 and 2007, 3.4% of screened pregnant women immigrants from Latin American countries were seropositive to *T. cruzi* [8]. Thus, efforts to control vector transmission in endemic countries is fundamental to reducing the burden of disease also in countries without triatomine vectors [9]. Current control methods include Indoor Residual Spraying (IRS) of insecticides to reduce domestic and peridomestic vector populations, maternal screening to detect and address congenital transmission, and blood donor screening [5, 10].

In the Southern Cone countries of South America, the main vector is *Triatoma infestans*. This species is predominantly endophilic and endophagic with widespread breeding colonies inside households and animal sheds; household infestations are particularly abundant in poorly constructed buildings in which wall and ceiling crevices provide triatomine refuges [11, 12]. The Southern Cone Initiative (INCOSUR), has promoted a coordinated international effort to combat domestic infestation by *Tri. infestans* and other domiciliated vectors using IRS [13, 14]. This resulted in substantial reductions in Chagas disease incidence and consequent certification by WHO of interrupted vectorial transmission in some countries (Uruguay, Chile, and some parts of Argentina, and Brazil) [10, 13].

Despite the successes of INCOSUR, vectorial transmission of *T. cruzi* persists in the American Gran Chaco, a seasonal dry forest ecosystem of 1.3 million km^2^ that straddles the borders of Bolivia, Argentina, and Paraguay [10]. The inhabitants in this region are some of the most marginalised, living in extreme poverty with little access to health care [15]. The incidence of *T. cruzi* infection and vectorial transmission amongst these communities are the highest worldwide [5, 16, 17, 18], and where26%-72% of houses are infested with *Tri. infestans* [11, 19], and 40%-56% of *Tri. infestans* are infected with *T. cruzi* [20, 21]. The majority (>93%) of all vectorial transmitted Chagas disease cases in the Southern Cone region occur in Bolivia [5].

IRS is currently the only widely deployed method to reduce human-*Tri. infestans* contact, being a historically proven strategy to reduce the burden of some vector-borne human diseases [22, 23]. The proportion of houses in a community with *Tri. infestans* infestation is one key measure used by health authorities to guide decision-making about IRS deployment, and, importantly, to justify the treatment of chronically infected children without the risk of reinfection [14, 24, 25, 26, 27]. Factors affecting IRS effectiveness, and the persistence of vectorial transmission in the Grand Chaco region, is variably attributed to poor building construction [17, 19], suboptimal IRS implementation practices and infestation surveillance methods [28], low public compliance to IRS requirements [29], short residuality of insecticide formulations [30, 31], and to *Tri. infestans* resistance and/or reduced susceptibility to insecticides [20, 32].

Synthetic pyrethroid insecticides are commonly used for IRS being lethal to susceptible triatomine populations. At low concentrations, pyrethroid insecticides are also used to flush out the vectors from wall crevices for purpose of surveillance [33]. Quality control studies of IRS practises are limited but indicate substantial variations in insecticide concentrations in sprayed houses, and frequently below the effective target concentration range [31, 34, 35, 36]. One reason for the lack of quality control studies is that high-performance liquid chromatography (HLPC), the gold standard method to measure insecticide concentrations, is technically challenging, costly, and generally not suited to endemic community settings. Recent advances in laboratory assays now provide alternative and relatively cheap methods to assess insecticide delivery and IRS practises [37, 38].

This study aimed to measure the variation in insecticide concentrations during routine IRS campaigns against *Tri. infestans* in the Bolivian Chaco. Insecticide concentrations were measured in prepared formulations in spray tanks, and in filter paper samples collected from sprayed houses. Factors potentially affecting delivery of insecticides to houses were also evaluated. In so doing, we adapted a chemical colorimetric assay to quantify pyrethroid concentrations in these samples.

## Methods

### Study site

This study was conducted in Itanambikua (20°1’5.94’’S; 63°30’41’’W) located in Camiri municipality, Santa Cruz Department, Bolivia (Fig. 1). The area forms part of the American Gran Chaco, characterized by seasonal dry forest with temperatures of 0°C-49°C, and rainfall of 500-1000mm/year [39]. Itanambikua is one of 19 ethnic Guarani communities in the municipality, holding a population of approximately 1,200 residents living in 220 houses which are constructed largely of sun-baked bricks (adobe), traditional wattle-and-daub (locally called “*tabique*”), wood, or a mixture of these materials. Additional buildings and structures near the house include animal sheds, storehouses, a kitchen and latrine, constructed of similar materials. The local economy is based on subsistence farming, mainly maize and peanuts, and small-scale breeding of poultry, pigs, goats, ducks and fish, the household surplus of which is sold in the local commercial town of Camiri (about 12 km distance).

**Figure 1.**
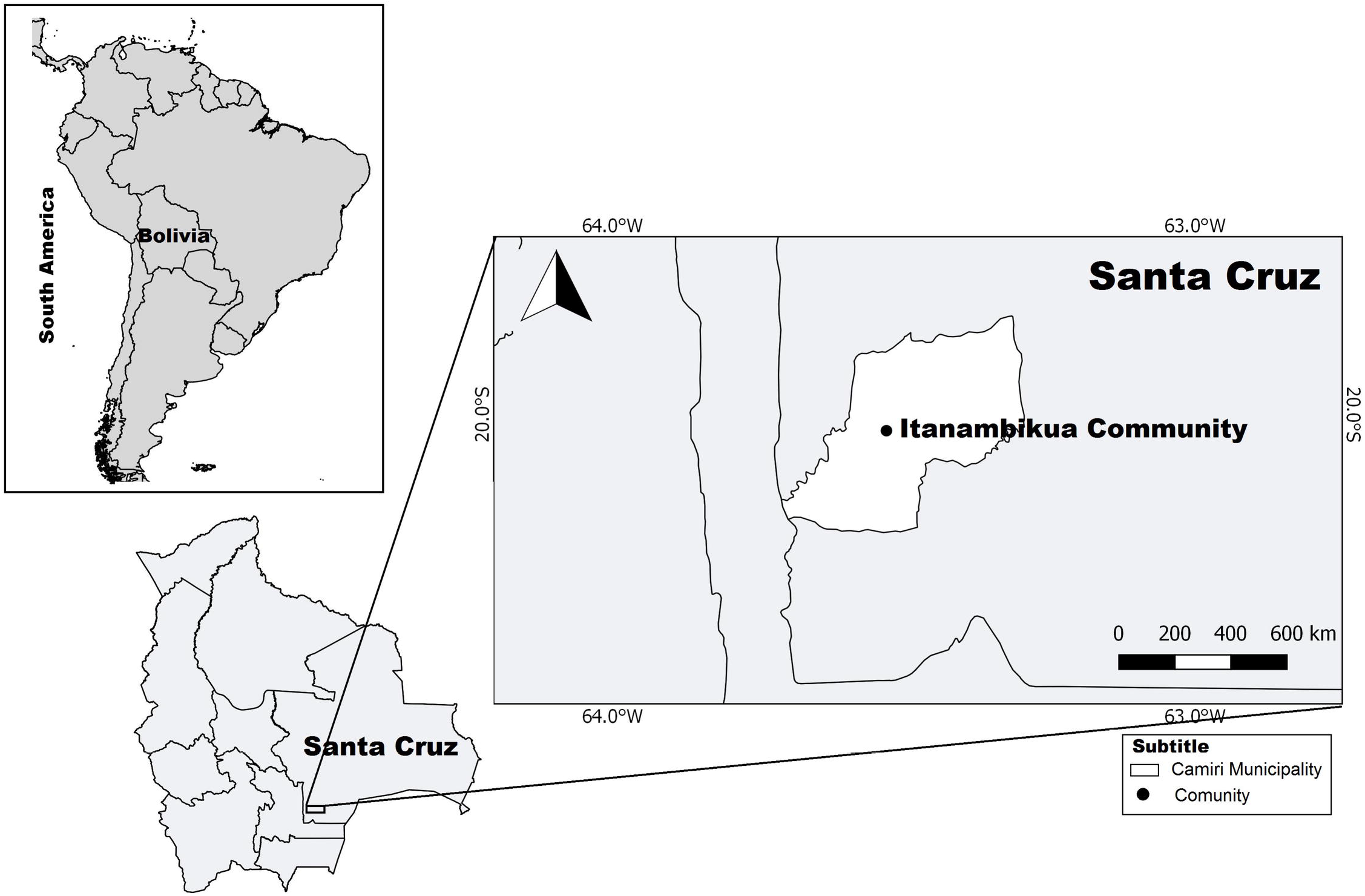
Location of the study site, Itanambikua community, located in Camiri Municipality, Santa Cruz department, Bolivia.

Camiri town also provides some employment to the community primarily in the building industry and domestic services.

During the current study, the prevalence of *T. cruzi* infection in children (2-15 year old) in Itanambikua was 20% [18]. Similar childhood infection seroprevalences are reported for nearby Guarani communities, showing an increase in prevalence with age with the vast majority of residents >30 years old infected [17]. Vectorial transmission is considered the main route of infection in these communities, the predominant vector being *Tri. infestans* which colonises houses and outbuildings [19, 20].

### Study design

#### IRS practises

In this community, IRS was performed by three resident trained health workers using alphacypermethrin [SC] 20% (Alphamost^®^, Hockley International Ltd., Manchester, United Kingdom). The insecticide was prepared at a delivery target concentration of 50mg active ingredient (a.i.)/m^2^ following the requirements of the Chagas Disease Control Program, Santa Cruz Administrative Department (*Servicio Departamental de Salud* - SEDES). The Insecticide was applied using a knapsack sprayer Guarany^®^ tank (Guarany Indústria e Comércio Ltda, Itu, São Paulo, Brazil), with 8L capacity, and equipped with a flat fan nozzle. The same health workers that mixed the spray tanks also sprayed the houses. These paid recruits had received training from the local municipality health authorities in insecticide preparation, and insecticide delivery to spray both the inside and outside walls of the house. They were also advised to request householders to empty their houses of all belongings including furniture (except for bed frames) the night before IRS was scheduled. The aim being to permit full access to the house interior for spraying. Compliance with this request was measured as outlined below.

#### Insecticide concentrations delivered to houses

To quantify the insecticide concentrations delivered to houses, researchers fitted filter papers (Whatman n°1; 55 mm diameter) to the wall surfaces of 57 houses just prior to IRS. Nine filter papers were fitted to each house divided between three wall heights (0.2, 1.2, and 2m above ground level), on each of three walls selected in an anti-clockwise direction starting from the main door. This provided three replicates at each wall height as recommended to monitor insecticide a.i. delivery [40]. Immediately after insecticide application, filter papers were collected by researchers and left to dry protected from direct sunlight. Once dry, filter papers were wrapped in Sellotape to protect and maintain the insecticide on the covered surface, and then wrapped in aluminium foil for storage at 7°C until testing. Of the total 513 filter papers collected, 480 from 57 houses were available for testing i.e. 8-9 filter papers per house. The tested samples included 437 filter papers from 52 adobe houses, and 43 filter papers from 5 *tabique* houses. This sample was in proportion to the relative abundance of house construction types in the community (76.2% [138/181] adobe, and 11.6% [21/181] *tabique*) as recorded by house-to-house survey as part of this study. The IQK™ assay for filter papers, and its validation against HPLC is described (Additional file 1). The target insecticide concentration range was 50mg a.i./m^2^ α 20% = 40-60mg a.i./m^2^.

#### Insecticide concentrations in the Guarany^®^ spray tank

Concentrations of insecticide a.i. in 29 spray tanks independently prepared by the health workers were quantified. Immediately after rigorous mixing of the formulation, 2ml of the solution was collected from the surface content. The 2ml sample was then vortexed for 5 minutes in the laboratory, and two 5.2μl sub-samples collected and tested as described below. The sedimentation rates of the insecticide a.i. in four prepared spray tanks were examined by random selection. Three sub-samples of 5.2μl were collected from the surface layer of each vortexed 2ml sample at intervals of one minute for 15 consecutive minutes post mixing. The IQK™ assay for spray tank solutions is described (Additional file 1). The target concentration tolerance range was 1.1-1.4mg a.i./ml (1.2mg a.i./ml ± 10%) based on the correct a.i. delivery onto filter papers as described above.

#### Assessment of spraying practises

To understand the relationship between insecticide spraying activities and insecticide delivery, one researcher (RG) accompanied two community IRS health workers during routine IRS, to 87 houses (30 houses in March 2016; 25 houses in November 2016; and 32 houses in January-February 2017). Details of the potential surface area inside and outside of houses to spray were measured, and the total time (minutes) that the health worker spent spraying was cryptically recorded. These raw data were used to calculate the spray rate defined as unit surface area sprayed per minute (m^2^/min). The recommended spray rate is 19m^2^/min [41]; the acceptable ± 10% spray rate tolerance range was 17.1-20.9m^2^/min.

Filter papers were fitted as described to the walls of 57 of these houses (25 sprayed in November 2016; and 32 in January-February 2017). To test whether the presence of filter papers influenced the health workers’ spraying activity, spray rates in 30 houses not fitted with filter papers (treated in March 2016), were compared to those in these 57 houses.

### Household’s compliance

Householders’ compliance to the request to empty their houses prior to IRS was recorded for 55 houses on a semi-quantitative scale of 0-2 (0 the majority of contents were left in the house; 1 most contents were removed; 2 houses were completely emptied). The influence of owner compliance on spray rates and delivery concentrations were examined.

### Statistical analyses

The insecticide concentrations on filter papers were examined by fitting to negative binomial Generalised Linear Models (GLMs) to test for variations between sprayed wall heights (3-levels); wall position (3-levels); spray rate (m^2^/min), date of IRS application, and the health worker’s identity (2-levels). All models including replicate samples from the same house included a cluster term (house ID) and other covariates as appropriate. Sedimentation of the insecticide a.i. concentrations in the spray tank solution over time were examined by including the starting value (time zero) as the model offset, and by inclusion of tank ID × time (days) interaction term. Analyses were performed using STATA v15.0 [42].

## Results

### Validation of IQK™ test

The accuracy of IQK™ chemistry to quantify alpha-cypermethrin concentrations was validated by comparing the values of 27 filter paper samples tested by IQK™ and HPLC, which showed a strong correlation (r=0.93; p<0.001) (Fig 2).

**Figure 2.**
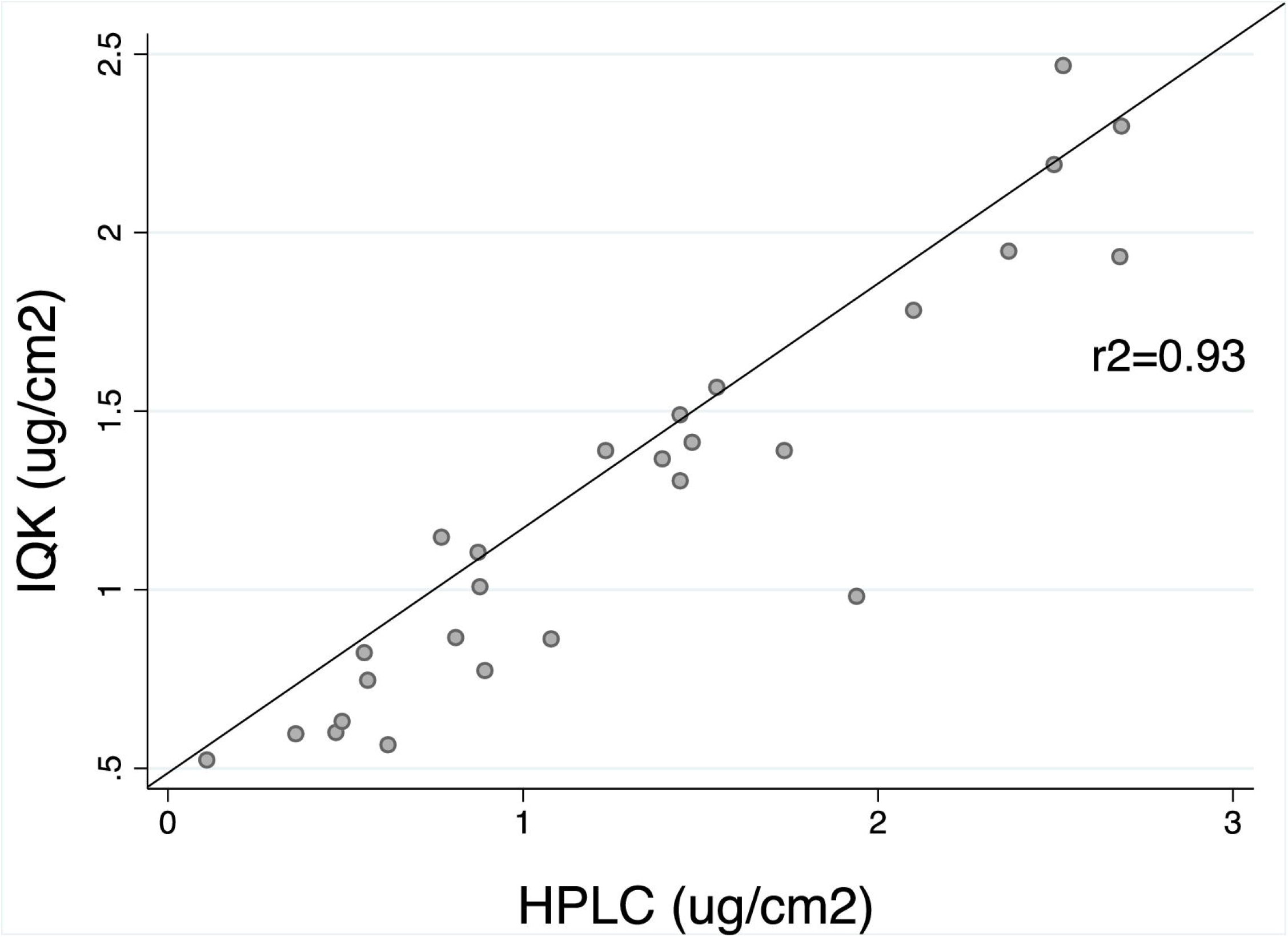
The association of the alpha-cypermethrin a.i. concentrations on 27 filter paper samples measured by HPLC and by IQK™ kit.

### Insecticide concentrations delivered to houses

A total 480 filter papers collected from 57 houses were tested by IQK™. Of these, only 10.4% (50/480) were within the target tolerance concentration of 40-60 mg a.i./m^2^ (Fig 3). The majority of the samples, 84.0% (403/480), were <40mg a.i./m^2^, and 5.6% (27/480) were >60mg a.i./m^2^. Median concentrations calculated for the 8-9 tested filter papers collected per house indicated that only 8.8% (5/57) of houses received the expected insecticide concentration; 89.5% (51/57) were lower, and 1.7% (1/57) were higher, than the target tolerance range (Fig 4).

**Figure 3.**
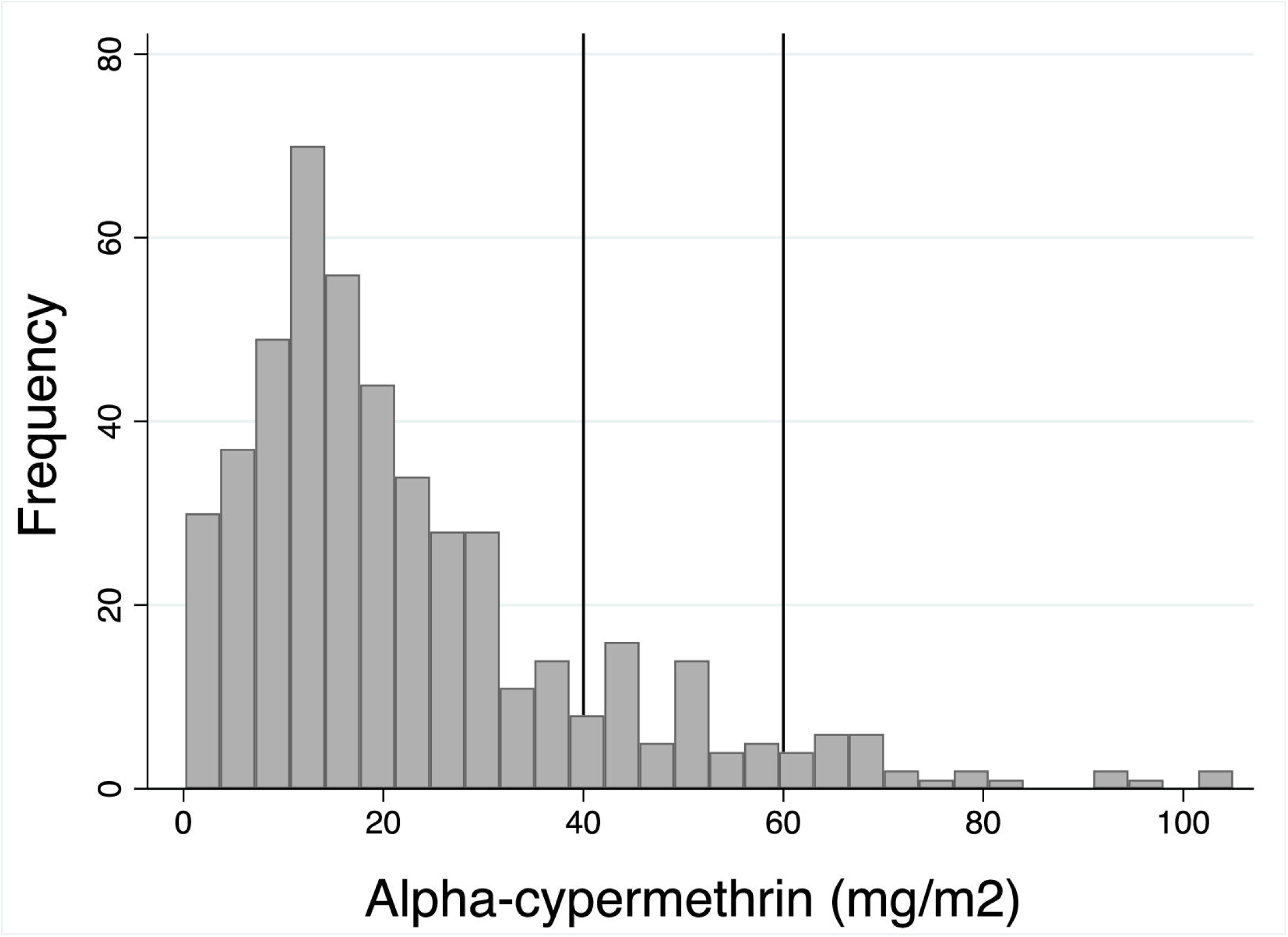
Frequency distribution of alpha-cypermethrin a.i. concentrations on filter paper fitted to 57 IRS-treated houses. The vertical lines represent the alpha-cypermethrin target a.i. concentration range (50mg α 20% a.i./m^2^).

**Figure 4.**
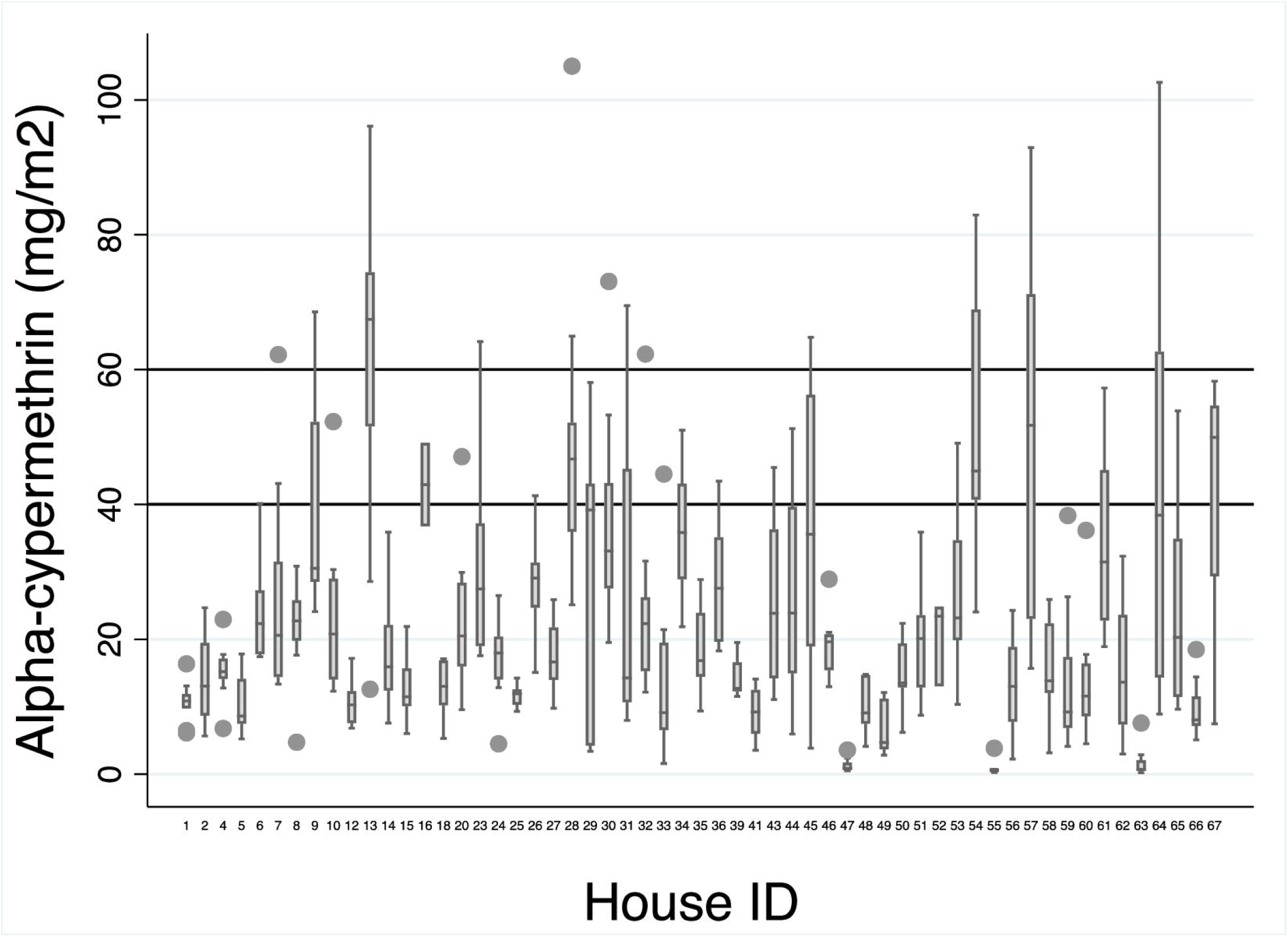
Median concentrations of alpha-cypermethrin a.i. delivered onto 8-9 filter papers per house in 57 IRS-treated houses. The horizontal lines represent the alpha-cypermethrin a.i. target concentration range (50mg α 20% a.i./m^2^). Error bars represents the lower and upper median adjacent values.

**Figure 5.**
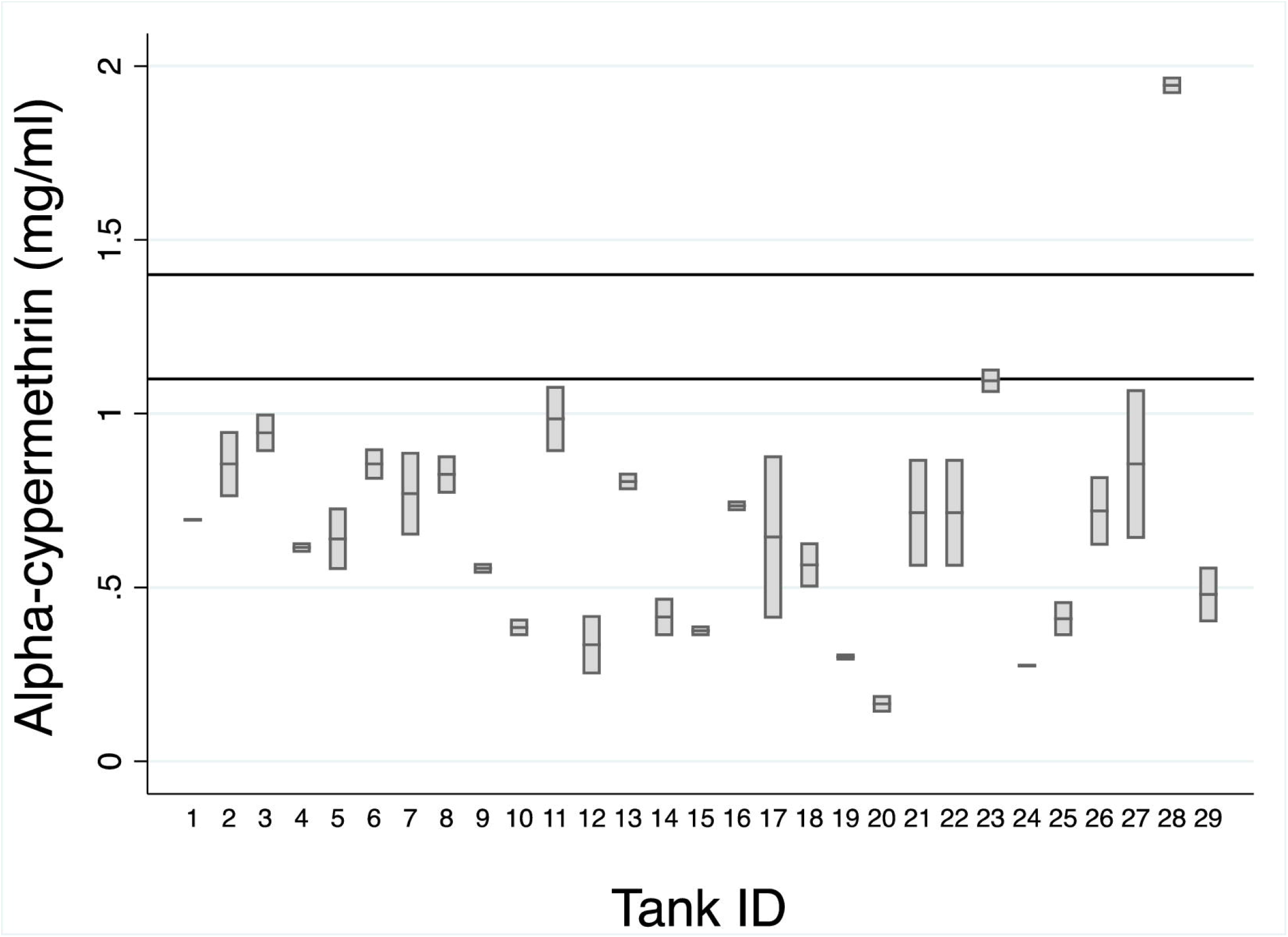
Median concentrations of alpha-cypermethrin a.i. in 29 spray tank preparations. Horizontal lines indicate the recommended a.i. concentrations (1.1-1.4mg/ml) to achieve the target a.i. concentration in houses of 40-60mg/m^2^.

The median concentrations delivered to filter papers at 0.2, 1.2 and 2.0m wall heights were 17.7mg a.i./m^2^ (IQR: 10.70-34.26), 17.3mg a.i./m^2^ (IQR: 11.43-26.91) and 17.6mg a.i./m^2^ (IQR: 10.85-31.37), respectively (Supplementary Figure S1). Controlling for wall ID and household spray rate, and clustering on house, the concentrations delivered at 1.2m height were marginally lower than those delivered at 2m (z=-2.46; p=0.014). Significant differences between other wall heights were not observed (z<1.85; p>0.10).

Accounting for this variation, significant differences in concentrations between wall positions were not detected (z<0.81; p>0.41). Neither were there significant differences associated with the date that IRS was conducted (z=1.80 p=0.07). The median concentrations delivered to the 5 *tabique* houses (18.8 mg/m^2^; IQR 11.76, 28.31), were not significantly different to those delivered to 52 adobe houses (23.17 mg/m^2^; IQR 13.86, 23.88) (z=0.13; p=0.89).

### Insecticide concentrations in spray tank preparations

The insecticide concentrations in the 29 independently prepared Guarany® spray tanks, sampled just prior to IRS application, varied by a magnitude of 12.1, from 0.16mg a.i./ml to 1.9mg a.i./ml per tank. Only 3.4% (1/29) spray tanks contained a.i. concentrations within the target tolerance range of 1.1-1.4mg a.i./ml; and 3.5% (1/29) tank were >1.4mg a.i./ml (Fig.5).

The median a.i. concentrations on filter papers per house in 21 houses, and those in the spray tanks used to spray the house, were not correlated (Spearman’s r^2^=-0.02; p=0.91); the relationship was not improved accounting for potential confounding variables (z=-0.25, p=0.8) (Fig 6).

**Figure 6.**
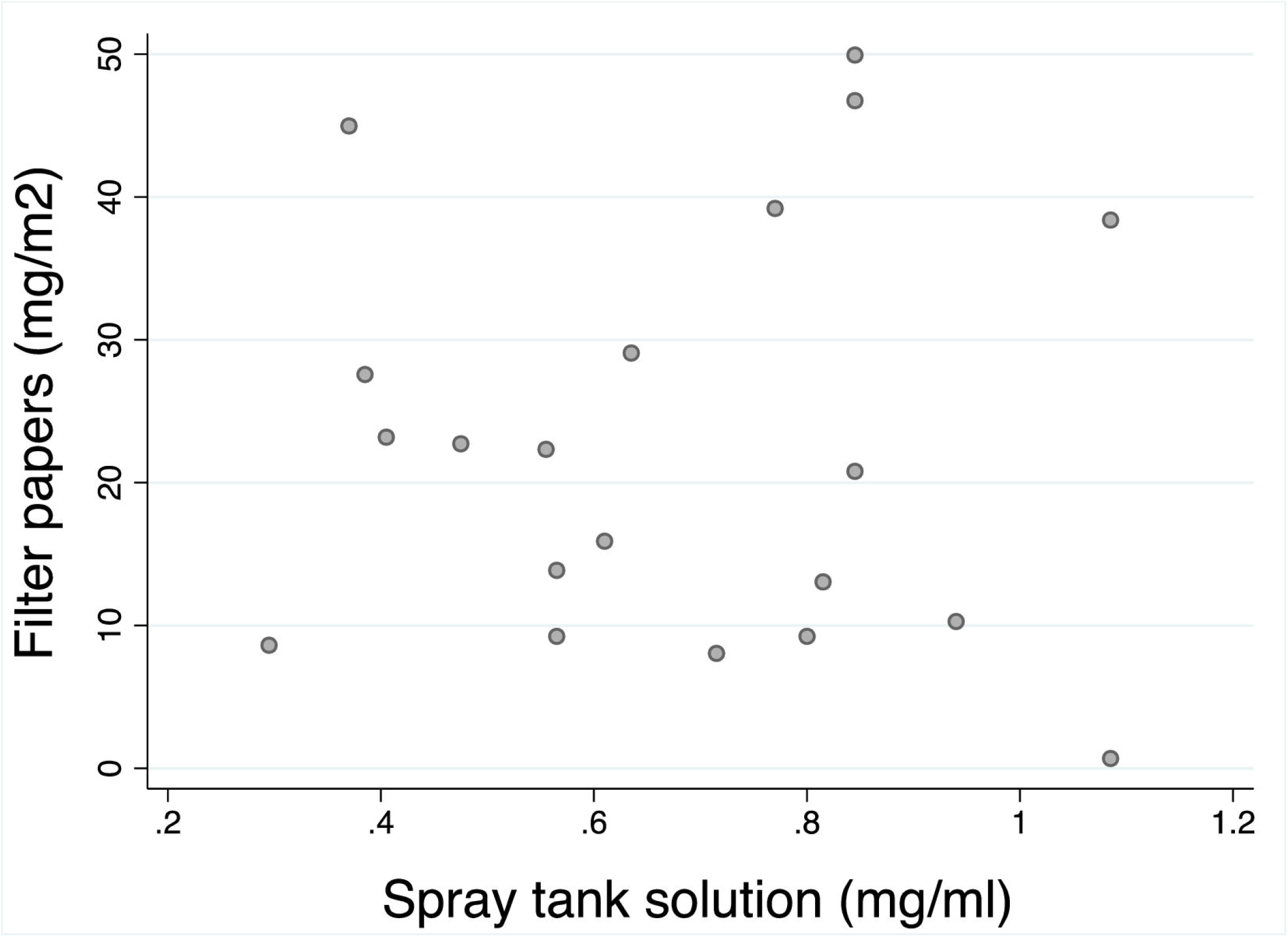
The association between the concentration of alpha-cypermethrin a.i. delivered on 8-9 filter papers per IRS-treated house, and in the prepared spray tank solutions used to treat each of the 21 houses.

### Sedimentation of the insecticide a.i. in the spray tank preparations

The insecticide concentrations in the surface solution of four spray tanks sampled immediately after vortexing (time 0 minutes) varied by a magnitude of 3.3 times (0.68-2.22 mg a.i./ml) (Kruskal-Wallis χ^2^_3_ =9.97; p=0.018) (Fig 7A). These values were within the target range for one tank, above target for one, and below target for the other two tanks. Thereafter, the insecticide a.i. concentrations in all four tanks significantly declined over the subsequent 14 minutes follow-up sampling (b=-0.04; z=-5.84; p<0.01); accounting for the individual tank starting values, the tank ID x time (minutes) interaction term was not significant (z<-0.03; p>0.54). Across the four tanks, the mean loss in insecticide a.i. was 29% (from 0.97 to 0.69mg a.i./ml) within 5 minutes, to 48.5% loss after 15 minutes (from 0.97 to 0.50mg a.i./ml) (Fig 7B).

**Figure 7.**
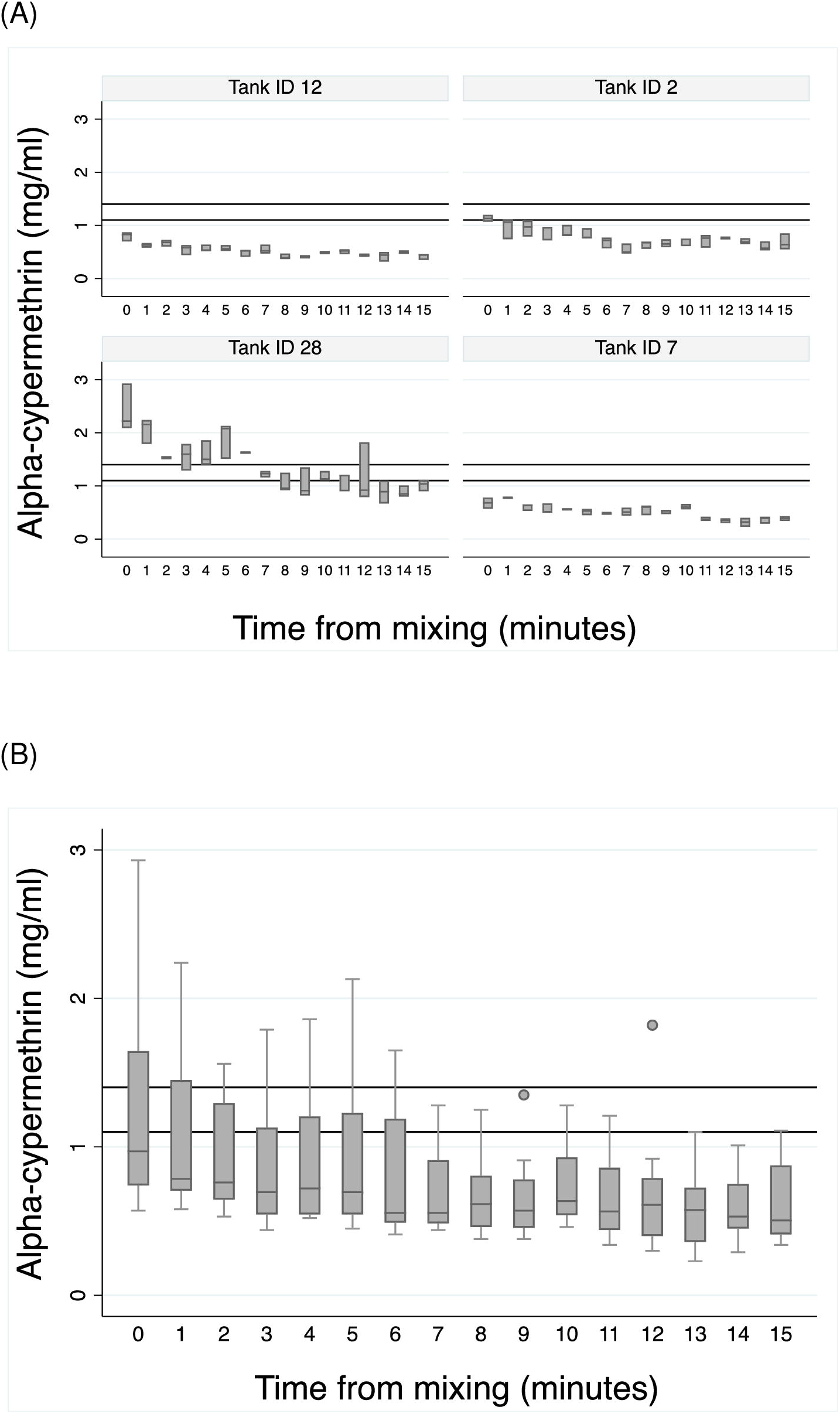
Sedimentation rates of alpha-cypermethrin a.i. in (A) four spray tanks measured each minute for 15 minutes from the time of mixing the tank contents and sampling. Values represent the median of three replicate sub-samples; (B) the median values of the four spray tanks in (A). Horizontal lines indicate the recommended a.i. concentrations (1.1-1.4mg/ml) to achieve the target a.i. concentration in houses of 40-60mg/m^2^. Error bars represents the lower and upper median adjacent values.

### Variations in spray rates

The wall surface area for potential IRS treatment was a median 128m^2^ (IQR: 99.0-210.0, range: 49.1-480.0) per house (n=87), and health workers spent a median 12 minutes (IQR: 8.2-17.5, range: 1.5-36.6) spraying each house. The time spent treating surfaces was positively associated with filter paper concentrations in the 56 houses that both variables were recorded (z=3.23; p=0.001), but the correlation was weak (Spearman’s r^2^=0.46) (Fig 8A). The median spray rate was 11 m^2^/min (IQR: 7.9-18.0, range: 3.0-72.7) (n=87 houses), was inversely related to insecticide concentrations delivered to filter papers (z=-4.99; p<0.001), however again the correlation was weak (Spearman’s r^2^=-0.5) (Fig 8B).

**Figure 8.**
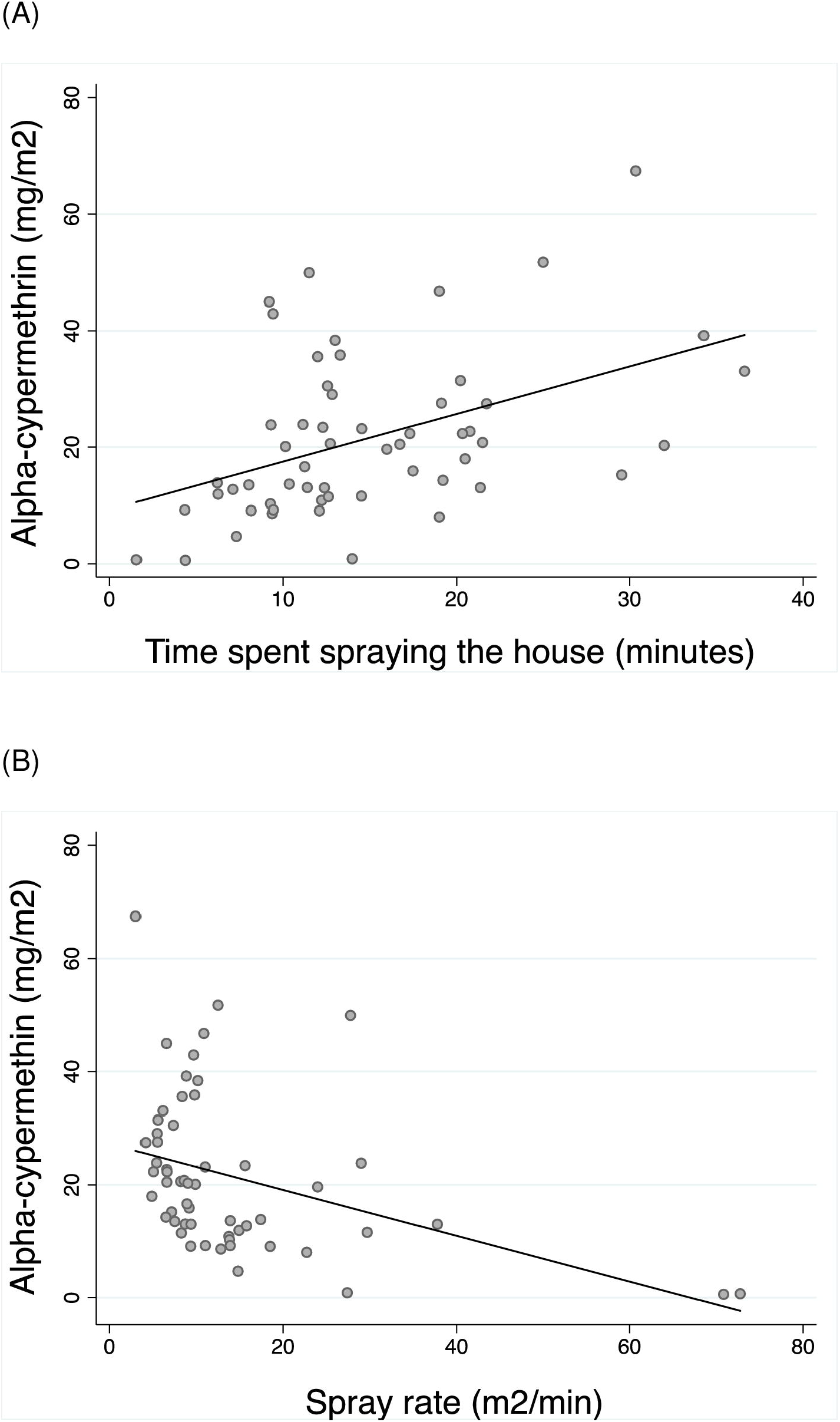
The association between median alpha-cypermethrin a.i. concentrations on 8-9 filter papers fitted to each of 56 IRS-treated houses, and (A) time spent spraying the house (minutes); (B) spray rate (m^2^/min).

The median spray rate in the 30 houses without fitted filter papers (15.8 m^2^/min, IQR 11.14, 22.5) was higher than in the 57 houses with fitted filter papers (9.35 m^2^/min, IQR 6.58, 14.79), but not statistical different when accounting for variations between health workers and household compliance (z=0.96; p=0.33). Different proportions of houses within these two groups were recorded with lower than expected spray rates <17.1 m^2^/min (53.3% [16/30] and 80.7% [46/57] of houses, respectively) (χ^2^_1_= 7.18; p=<0.007). Overall, only 6.9% (6/87) of houses were treated within the expected spray rate range; 71.3% (62/87) were lower (<17.1m^2^/min), and 21.8% (19/87) were higher (>20.9m^2^/min).

Only one of the health workers sprayed houses with filter papers fitted. Comparison of the variation in spray rates by this operator in houses with (n=57) and without (n=14) filter papers fitted, were not statistically different (Kruskal-Wallis χ^2^_1_ =0.65; p=0.41). In the absence of filter papers in 30 houses, the spray rates differed between the two observed health workers by 1.38 times (13.2m^2^/ min [IQR: 7.1-18.16] *versus* 19.3m^2^/ min [IQR: 15.1-35.6]) (z=3.09, p=0.002).

### Variations associated with householder compliance

Householder compliance to the request to empty their house in preparation for IRS varied: 31% (17/55) did not empty the house; 42% (23/55) semi-emptied the house (limiting wall access), and 27% (15/55) completely emptied the house.

Accounting for the variation between health workers, and the presence or absence of filter papers, spray rates in fully emptied houses were lower than in both non-emptied houses (z=6.76; p>0.001), and in semi-emptied houses (z=3.53; p>0.001). The absolute time spent spraying houses in these three groups were not dissimilar (z<-0.05; p>0.16), whereas the houses sizes were different: the median surface area to spray in houses fully emptied (104m^2^ [IQR: 60.0-169.0m^2^], range: 49-320m^2^) was statistically smaller than in houses of the other two categories (non-emptied: 224m^2^ [IQR: 168.0-280.0m^2^], range: 100-420m^2^; semi-emptied: 132m^2^ [IQR: 108.0-384.0m^2^], range: 60-480m^2^) (z>2.59; p<0.01). The higher spray rates in non-emptied houses (18.7m^2^/min, IQR: 13.3-37.9) were closer to the WHO recommended spray rates than the spray rates of the other houses (10.2m^2^/min, IQR: 6.58-15.39).

## Discussion

Successes in reducing Chagas disease burdens in the Americas is attributed, in part, to IRS vector control campaigns [10, 43, 44, 45]. In contrast, IRS activities against *Tri. infestans* vectors in the Gran Chaco region have not been noticeably successful evidenced by the high incidence of vector-borne transmission [16, 17, 18, 28, 29]. This study evaluated IRS practises and procedures in Itanambikua, identified as one such focus of chronic vector-borne transmission [18]. Although the study focuses on a single community, it appeared to be similar to other rural communities in the Bolivian Chaco, with similarities in childhood infection rates [16, 17, 18], conditions of the indigenous (Guarani) houses, persistent of domestic infestations and concerns about the vector control activities [17, 19].

This study revealed that the alpha-cypermethrin delivered during routine IRS varied substantially between and within sprayed houses, and that most filter papers and houses (84%-89.5%) received less than the target concentration. The observed variation in a.i. concentrations and suboptimal delivery is not unique. In India, only 7.3% (41/560) of sprayed houses received DDT at target concentrations, with similarly large variations within and between houses [35]. In Nepal, the average filter paper received 1.74mg a.i./m^2^ (range: 0.0-17.5 mg/m^2^) which was only 7% of the target concentration (25mg a.i./m^2^) [36]. HPLC analysis of filter papers revealed extensive variations in deltamethrin a.i. of 12.8-51.2mg a.i./m^2^ delivered to walls, and 4.6-61.0mg a.i./m^2^ delivered to roofs of Paraguayan Chaco houses [31], and in Tupiza, Bolivia, the Chagas control program reported deltamethrin concentrations of 0.0-59.6 mg/m^2^ delivered to 5 houses monitored by HPLC [34].

Campaigns to control other vectors have been able to achieve high quality control. For example, in Bioko Island, Republic of Equatorial Guinea, HPLC analysis of IRS conducted using Primiphos-methyl collected onto glue dots reported that 82% (49/60) of houses received the recommended concentrations of 5g/m^2^ [46]. And in Vanatu Island, South West Pacific, 83% (27/30) of houses sprayed with lambda-cyhalothrin received the expected concentrations based on IQK™ [37].

A potential contributing factor to the incorrect delivery of insecticide to houses is the initial concentrations in the prepared spray tanks prior to spraying. In the current study, observation of the health workers by the researchers confirmed that they followed the insecticide preparation formulae, and training to vigorously mix the solutions once diluted in the spray tank. Nonetheless, analysis of the tanks’ contents demonstrated a.i. concentration that varied twelve-fold; furthermore, only 3.4% (1/29) of tank solutions tested were under the acceptable lower concentration limit of 1.1 mg a.i./ml which was required to achieve the target delivery concentration.

Inappropriate preparation of insecticide formulations has been reported in other vector control programs. For example, in the Indian leishmaniasis control program, of 51 monitored spray teams, only 29% prepared and mixed DDT solutions correctly, and none filled the spray tanks according to guidelines [47]. Assessment in Bangladesh villages showed similar trends: only 42-43% of district IRS teams prepared the insecticide and filled the spray tanks according to protocol; in one district this value was only 7.7% [48].

To investigate further the variation in insecticide concentrations in the spray tanks, we observed high rates of a.i. sedimentation in laboratory simulations. However, we found no correlation between the concentrations in spray tank preparations and that delivered onto filter papers in matched houses. Sedimentation of insecticide a.i. is described as an important factor that affects IRS quality e.g. for DDT formulated as water-dispersible powders [35, 49]. Suspension concentrates [SC], is widely used for IRS against *Tri. infestans*, and one insecticide formulation applied by SEDES. As for all insecticide formulations, they are prone to sedimentation during the dilution process, even after rigorous mixing, the physical stability depending on many factors particularly the a.i. compound particle size, and other compounds used in their formulation. Methods to control the physical stability of SC formulations is under investigation [50]. Notwithstanding, SC formulations are successfully deployed to reduce domestic infestations of *Tri. infestans* in other regions of Latin America [51].

The volume of insecticide released from the spray tank is dependent of the spray nozzle specifications, the pressure inside the tank, and time spent spraying the target surface [41]. The recommended time that health workers should spend spraying houses using a flat spray nozzle (discharging 757ml/minute at standard tank pressure of 280kPa), is approximately 19m^2^/minute [41]. The recorded spray rate in the current study ranged from 3.0 to 72.7 m^2^/min, and only 6.9% (6/87) of houses were treated within the acceptable spray rate range of 17.1 to 20.9 m^2^/min (i.e. 19m^2^/min ± 10%). The delivered concentrations on filter papers were poorly correlated with the recorded spray rates, indicating that additional factors are involved in generating the variation in delivered concentrations.

Health workers showed differences in median spray rates, and in delivery to different wall heights, suggesting that the spraying motion down a wall was not consistent. The locally recruited health workers (residents of the Guarani communities) in this study received a single day training course by the municipality public health authority, including basic instructions in insecticide preparation and delivery using a Guarany® sprayer, and were provided with the insecticide. The authors did not identify any training manual, local or national IRS guidelines, or community records of annual IRS coverage.

The health workers were also recommended to request householders to empty their houses of all belongings including heavy furniture, in order to facilitate access for IRS delivery. Compliance to this request is documented to affect the quality and insecticide coverage of houses [52]. In this study, compliance was variable with only 27% of houses being fully emptied, and 31% were not emptied. Lower spray rates were observed in houses that were fully emptied or partially emptied compared to those that were not emptied. Since health workers spent similar amounts of time spraying houses across these levels of compliance, the difference in spray rates is partially related to the wall surface area available to spray. Houses that were fully emptied were statistically smaller than those in the other two compliance categories, hence greater compliance was seen in smaller houses, presumably because they are easier to empty. It is possible that the larger non-emptied houses were quicker to finish spraying due to the restricted access, hence the apparent higher spray rates in these houses, which more closely approximating the WHO recommended spray rates, but at the expense of smaller areas of insecticide coverage. Such distinctions between house sizes and levels of compliance may prove immaterial since we did not detect statistical differences in the delivered insecticide concentrations between these compliance categories, though we acknowledge that we did not adjust the calculated total surface area to spray according to specific measurements of obstacles (e.g furniture) not removed from the house. Further investigation would be informative.

In this study, we used the recently developed IQK™ chemistry to estimate alpha-cypermethrin a.i. concentrations. Previous studies have used IQK™ to quantify pyrethroids, bendiocarb and DDT, to quantify delivered and residual concentrations on a variety of sampling mediums including adhesive tape, felt pads, glue dots, long-lasting insecticidal nets (LLINs), and in scrapings from treated wall surfaces [37, 49, 53, 54, 55]. Unlike the previous methods, here we adapted the IQK™ assay conditions to quantify alpha-cypermethrin a.i. captured onto filter papers and in tank solution samples. We demonstrate that the values generated by IQK™ were strongly correlated (r^2^=0.93) with those estimated using HPLC analysis, the gold standard method, thus further validating the assay accuracy. A similarly high correlation was reported between estimates using IQK™ kit bench-marked against HPLC during evaluation of residual deltamethrin concentrations in LLINs [55]. Consequently, it is not likely that the observed variation in the insecticide concentrations in this study were due to inaccuracy of the quantification methods. HPLC analysis is relatively expensive and requires specialist equipment and a high level of staff training, thus is not usually suited to endemic settings. By contrast, IQK™ is low-tech, providing readings within 30 minutes, and cost-effective (< $ 10 per assay), making it a useful tool to locally support quality control, training, and decision making regarding equipment performance e.g. [38, 53]

WHO guidelines to measure insecticide delivery are to locate at least 4 filter papers on different walls and at different wall heights prior to spraying [40]. The use of filter papers for insecticide capture is a logistically easy method, but being visible to spray teams, potentially could influence their performance. In this study, no significant difference was detected in spray rates in houses without filter papers (15.8 m^2^/min) compared to those with filter papers fitted (9.35 m^2^/min), suggesting that the presence of filter papers did not influenced health worker behaviour. Furthermore, filter papers or other a.i. capturing materials such as small felt pads [37] could be designed to be less conspicuous.

### Implications of Chagas disease control

The consequences of poor vector control are far reaching. Bolivian Chaco communities register childhood infection prevalences of 20%-25%, with annual force of infection rates of 0.021 and 0.046 [16, 17, 18]. Following Bolivian MoH guidelines, treatment of chronic infected children (children under 15 years-old, positive for *T. cruzi* antibodies) is advised only once communities are under successful vector control, defined as ≤3% of the houses in the community being infested, and proven absence of triatomine nymphal stages in the patient’s house [27]. Thus, confirmation of a successful vector control programme also relies on accurate triatomine surveillance methods which currently is based on timed manual capture [56], but which is criticised for its low sensitivity [57], particularly to detect infestations in wall crevices [58]. Residual house infestation post-IRS is commonly observed in the Gran Chaco region [59, 60, 61]. The longer term consequences of successive suboptimal exposure to insecticides can lead to genotype selection in the vector population promoting insecticide resistance or tolerance [62], as already reported in the Chaco region [20, 63].

## Conclusions

IRS is the only vector control activity against *Tri. infestans* in the Chaco region. The collective result of this study demonstrate the generally known abiotic and biotic complexities to achieve effective and sustainable vector control. The suboptimal concentrations observed here were partially attributed to the insecticide physical characteristics, but also indicate the need for revision of IRS practises including training of health workers and community education to encourage compliance. The quality of IRS delivery and coverage will have important bearings on the residuality of insecticidal effectiveness in houses, and thus on the interval between required IRS campaigns. Strategies to improve the rigour of IRS practises in the region are possible [64], and IQK™ is an available tool to facilitate the much needed changes.

## Supporting information

Additional file 1 laboratory assays

## Data Availability

The data supporting the conclusions of this article are included within the article. Data are available from the corresponding authors on reasonable request to be used solely within the context of this study, following the ethical agreements, and with permission from the relevant authorities and co-authors.

## Supplementary S1 Figures

**Figure S1.**
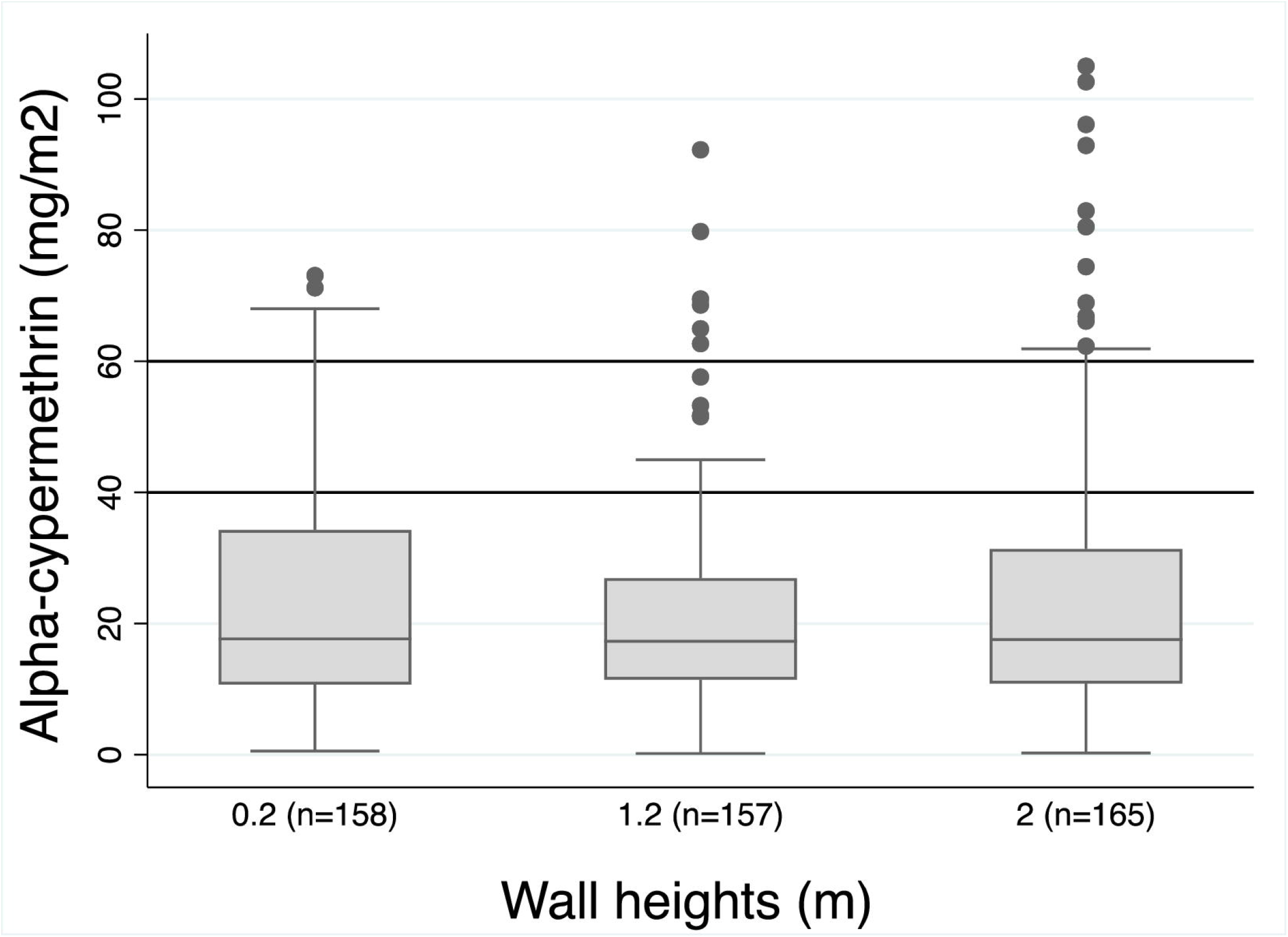
Median alpha-cypermethrin a.i. concentrations on filter papers fitted to houses at different wall heights. The horizontal lines represent the alpha-cypermethrin a.i. target concentration range (50mg α 20% a.i./m^2^). Error bars represents the lower and upper median adjacent values.

## Declarations

### Ethics approval and consent to participate

This study did not require human or animal ethical approval.

The IRS campaigns were carried out by the local health workers as routine and as directed by the Chagas Disease Control Program, Santa Cruz Administrative Department (*Servicio Departamental de Salud*-SEDES). Permission to accompany the IRS health workers to record their practises was authorized by SEDES, and by the health authorities at provincial (*Red de Salud Cordillera*, Santa Cruz) and municipal (Secretary of Human development, Camiri) levels, by the Guarani Indigenous District office (*Kaami Capitania*), and by the local community leader and steering committee (Itanambikua community). The purpose of the observing researcher’s presence was fully explained to householders who gave their verbal consent.

### Consent for publication

Not applicable

### Authors’ contributions

OC, CB and RG designed the study. RG collected field samples and complied the data. RG, RAEL HMI performed the laboratory assays. RG, OC and MJIP conducted data analysis. RG and OC drafted the manuscript and performed the statistical analyses. All authors contributed to the draft review and approved the final manuscript.

## Acknowledgements

We thank the Itanambikua community leader E. Arteaga, steering committee and community health workers P. Rua, L. Rua, O. Robles, and field assistants P. Jardilo and E. Rua, for their technical support. We also acknowledge H. Bettinsoli (manager of Red de Salud Cordillera, Santa Cruz), the Guarani Kaami Capitania, R. Vargas (manager of Chagas disease Control Program Santa Cruz Administrative Department-SEDES), and M. Calderón and her team (Camiri municipality health office), for their guidance. We thank F. Tinajeros (John Hopkins University) for assistance with logistics, and a special thanks to E. Liendo (Ministerio de Salud, Bolivia, and community medical doctor) for field support, and to the residents for their willingness to participate in the study.

## Additional file 1

File name: Additional file 1

File format: docx

Title of data: Additional file 1

Description of data: Protocol of laboratory assays

## References

1. Bern C. Chagas’ Disease. 2015. New England Journal of Medicine 373 (5): 456–466. doi: 10.1056/NEJMra1410150.

2. Kyu HH, Abate D, Abate KH, Abay SM, Abbafati C, Abbasi N, et al. Global, regional, and national disability-adjusted life-years (DALYs) for 359 diseases and injuries and healthy life expectancy (HALE) for 195 countries and territories, 1990-2017: a systematic analysis for the Global Burden of Disease Study 2017. 2018. doi:10.1016/S0140-6736(18)32335-3

3. Roberts NLS, Mountjoy-Venning WC, Anjomshoa M, Banoub JAM, Yasin YJ. GBD 2017 Disease and Injury Incidence and Prevalence Collaborators. Global, regional, and national incidence, prevalence, and years lived with disability for 354 diseases and injuries for 195 countries and territories, 1990-2017: a systematic analysis for the Global Burden of Disease Study (vol 392, pg 1789, 2018). Lancet. 2018;393 doi:10.1016/S0140-6736(18)32279-7.

4. Yasin YJ, Banoub JAM, Husseini A. GBD 2017 Causes of Death Collaborators. Global, regional, and national age-sex-specific mortality for 282 causes of death in 195 countries and territories, 1980-2017: a systematic analysis for the Global Burden of Disease Study 2017 (vol 392, pg 1736, 2018). Lancet. 2018. doi:10.1016/S0140-6736(18)32203-7.

5. World Health Organization. Chagas disease in Latin America: an epidemiological update based on 2010 estimates. http://www.who.int/wer (2015). 90.Access: 28/09/2020.

6. Pinazo MJ, Munoz J, Posada E, Lopez-Chejade P, Gallego M, Ayala E, et al. Tolerance of Benznidazole in Treatment of Chagas’ Disease in Adults. Antimicrobial Agents and Chemotherapy. 2010;54 11:4896–9; doi: 10.1128/aac.00537-10. WOS:000284155000051.

7. Lee BY, Bacon KM, Bottazzi ME, Hotez PJ. Global economic burden of Chagas disease: a computational simulation model. Lancet Infectious Diseases 2013; 13 (4): 342–348. doi: 10.1016/s1473-3099(13)70002-1.

8. Munoz J, Coll O, Juncosa T, Verges M, del Pino M, Fumado V, et al. Prevalence and Vertical Transmission of Trypanosoma cruzi Infection among Pregnant Latin American Women Attending 2 Maternity Clinics in Barcelona, Spain. Clinical Infectious Diseases. 2009;48 12:1736–40; doi: 10.1086/599223. WOS:000266439100017.

9. Liu Q, Zhou XN. Preventing the transmission of American trypanosomiasis and its spread into non-endemic countries. Infectious Diseases of Poverty. 2015;4:11; doi: 10.1186/s40249-015-0092-7. WOS:000370025600001.

10. Schofield CJ, Jannin J, Salvatella R. The future of Chagas disease control. Trends Parasitol. 2006;22 12:583–8; doi: 10.1016/j.pt.2006.09.011.

11. Gurevitz JM, Ceballos LA, Sol Gaspe M, Alvarado-Otegui JA, Enriquez GF, Kitron U, et al. Factors Affecting Infestation by Triatoma infestans in a Rural Area of the Humid Chaco in Argentina: A Multi-Model Inference Approach. Plos Neglected Tropical Diseases. 2011;5 10; doi: 10.1371/journal.pntd.0001349. WOS:000296579700022.

12. Gürtler RE, Yadon ZE. Eco-bio-social research on community-based approaches for Chagas disease vector control in Latin America. Trans R Soc Trop Med Hyg 2015; 109 (2): 91–98. doi: 10.1093/trstmh/tru203.2015.

13. Dias JCP. Southern Cone Initiative for the elimination of domestic populations of Triatoma infestans and the interruption of transfusional Chagas disease. Historical aspects, present situation, and perspectives. Memorias Do Instituto Oswaldo Cruz 2007; 102 11–18. doi: 10.1590/s0074-02762007005000092.

14. Dias JCP, Schofield CJ. The evolution of Chagas disease (American trypanosomiasis) control after 90 years since Carlos Chagas discovery. Memorias Do Instituto Oswaldo Cruz 1999; 94 103–121. doi: 10.1590/s0074-02761999000700011.

15. Argentina. Ministério de Hacienda. Secretaria de Hacienda. Dirección Nacional de Asuntos Provinciales: Chaco. Informe sintético de caracterización socio-productiva 20p. http://www2.mecon.gov.ar/hacienda/dinrep/Informes/archivos/chaco.pdf (2018). Accessed 03/09/2020.

16. Chippaux J-P, Postigo JR, Santalla JA, Schneider D, Brutus L. Epidemiological evaluation of Chagas disease in a rural area of southern Bolivia. Transactions of the Royal Society of Tropical Medicine and Hygiene 2008; 102 (6): 578–584. doi: 10.1016/j.trstmh.2008.03.008.

17. Samuels AM, Clark EH, Galdos-Cardenas G, Wiegand RE, Ferrufino L, Menacho S, et al. Epidemiology of and Impact of Insecticide Spraying on Chagas Disease in Communities in the Bolivian Chaco. Plos Neglected Tropical Diseases 2013; 7 (8): e2358. doi: 10.1371/journal.pntd.0002358.

18. Hopkins T, Goncalves R, Mamani J, Courtenay O, Bern C. Chagas disease in the Bolivian Chaco: Persistent transmission indicated by childhood seroscreening study. International Journal of Infectious Diseases 2019; 86 175–177. doi: 10.1016/j.ijid.2019.07.020.

19. Lardeux F, Depickere S, Aliaga C, Chavez T, Zambrana L. Experimental control of Triatoma infestans in poor rural villages of Bolivia through community participation. Transactions of the Royal Society of Tropical Medicine and Hygiene 2015; 109 (2): 150–158. doi: 10.1093/trstmh/tru205.

20. Santo-Orihuela PL, Vassena CV, Carvajal G, Clark E, Menacho S, Bozo R, et al. Toxicological, Enzymatic, and Molecular Assessment of the Insecticide Susceptibility Profile of Triatoma infestans (Hemiptera: Reduviidae, Triatominae) Populations From Rural Communities of Santa Cruz, Bolivia. Journal of Medical Entomology. 2017;54 1:187–95; doi: 10.1093/jme/tjw163. WOS:000397207700025.

21. Perez E, Monje M, Chang B, Buitrago R, Parrado R, Barnabe C, et al. Predominance of hybrid discrete typing units of Trypanosoma cruzi in domestic Triatoma infestans from the Bolivian Gran Chaco region. Infection Genetics and Evolution 2013: 13 116–123. doi: 10.1016/j.meegid.2012.09.014.

22. Wilson AL, Courtenay O, Kelly-Hope LA, Scott TW, Takken W, Torr SJ, et al. The importance of vector control for the control and elimination of vector-borne diseases. Plos Neglected Tropical Diseases 2020; 14 (1): e0007831. doi: 10.1371/journal.pntd.0007831.

23. World Health Organization. Global vector control response 2017-2030. 51 p. https://www.paho.org/en/documents/global-vector-control-response-2017-2030 (2017).

24. Dias JCP. Control of Chagas disease in Brazil. Parasitology Today. 1987;3 11:336–41; doi: 10.1016/0169-4758(87)90117-7.

25. Aiga H, Sasagawa E, Hashimoto K, Nakamura J, Zuniga C, Chevez JER, et al. Chagas Disease: Assessing the Existence of a Threshold for Bug Infestation Rate. American Journal of Tropical Medicine and Hygiene. 2012;86 6:972–9; doi: 10.4269/ajtmh.2012.11-0652. WOS:000304785700012.

26. PAHO. Pan-American Health Organization. Taller para el Establecimiento de Pautas Tecnicas en el Control de Triatoma dimidiata. https://www.paho.org/uru/index.php?option=com_docman&view=download&alias=58-taller-para-el-establecimiento-de-pautas-tecnicas-en-el-control-de-tiatoma-dimidiata&category_slug=manuales-y-guias&Itemid=307 (2002). Accessed 03/09/2020.

27. Bolivia. Ministerio de Salud y Deportes Unidad de Epidemiologia, Programa Nacional de Chagas.: Manual de procesos para la detección, diagnóstico, tratamiento y seguimiento de la enfermedad de Chagas infantil. 99p. https://www.minsalud.gob.bo/images/Documentacion/dgss/Epidemiologia/NORMATIVOS%20PNCH/Manual%20de%20Procesos%2031.pdf (2007). Accessed 03/09/2020.

28. Gurtler RE. Sustainability of vector control strategies in the Gran Chaco Region: current challenges and possible approaches. Memorias Do Instituto Oswaldo Cruz. 2009;104:52–9. WOS:000269123500008.

29. Gurtler RE, Kitron U, Cecere MC, Segura EL, Cohen JE. Sustainable vector control and management of Chagas disease in the Gran Chaco, Argentina. Proceedings of the National Academy of Sciences of the United States of America. 2007;104 41:16194–9; doi: 10.1073/pnas.0700863104. WOS:000250128800041.

30. Rojas de Arias A, Lehane MJ, Schofield CJ, Fournet A. Comparative evaluation of pyrethroid insecticide formulations against Triatoma infestans (Klug): residual efficacy on four substrates. Mem Inst Oswaldo Cruz. 2003;98 7:975–80. doi: 10.1590/s0074-02762003000700020.

31. Rojas de Arias A, Lehane MJ, Schofield CJ, Maldonado M. Pyrethroid insecticide evaluation on different house structures in a Chagas disease endemic area of the Paraguayan Chaco. Mem Inst Oswaldo Cruz. 2004;99 6:657–62; doi: /S0074-02762004000600022.

32. Picollo MI, Vassena C, Orihuela PS, Barrios S, Zaidemberg M, Zerba E. High resistance to pyrethroid insecticides associated with ineffective field treatments in Triatoma infestans (Hemiptera : Reduviidae) from northern Argentina. Journal of Medical Entomology. 2005;42 4:637–42; doi: 10.1603/0022-2585(2005)042[0637:hrtpia]2.0.co;2. WOS:000230406400017.

33. Zerba EN. Susceptibility and resistance to insecticides of Chagas disease vectors. Medicina-Buenos Aires. 1999;59:41–6.

34. Guillen G, Diaz R, Jemio A, Cassab JA, Pinto CT, Schofield CJ. Chagas disease vector control in Tupiza, southern Bolivia. 1997. <Go to WOS:A1997WE10000001.

35. Coleman M, Foster GM, Deb R, Singh RP, Ismail HM, Shivam P, et al. DDT-based indoor residual spraying suboptimal for visceral leishmaniasis elimination in India. Proceedings of the National Academy of Sciences of the United States of America 2015. 112 (28): 8573–8578. doi: 10.1073/pnas.1507782112.

36. Chowdhury R, Huda MM, Kumar V, Das P, Joshi AB, Banjara MR, et al. The Indian and Nepalese programmes of indoor residual spraying for the elimination of visceral leishmaniasis: performance and effectiveness. Annals of Tropical Medicine and Parasitology 2011; 105 (1): 31–45. doi: 10.1179/136485911x12899838683124.

37. Russell TL, Morgan JC, Ismail H, Kaur H, Eggelte T, Oladepo F, et al. Evaluating the feasibility of using insecticide quantification kits (IQK) for estimating cyanopyrethroid levels for indoor residual spraying in Vanuatu. Malaria Journal. 2014;13; doi: 10.1186/1475-2875-13-178. WOS:000336630900001.

38. Kaur H, Eggelte T: In Colorimetric Assay for Pyrethroid Insecticides, Vol. WO/2009/106845, G01N 31/22 (2006.01) edition. Edited by World Intellectual Property Organization; 2009. - In Colorimetric assay. 2009

39. Pennington RT, Prado DE, Pendry CA. Neotropical seasonally dry forests and Quaternary vegetation changes. Journal of Biogeography. 2000;27 2:261–73. WOS:000089193500003.

40. World Health Organization: Guidelines for testing mosquito adulticides for indoor residual spraying treatment of mosquito nets. 60p. https://apps.who.int/iris/bitstream/handle/10665/69296/WHO_CDS_NTD_WHOPES_GCDPP_2006.3_eng.pdf?sequence=1&isAllowed=y (2006). Access: 28/0/2020.

41. World Health Organization Vector Control: Methods for use by individuals and communities. 412 p. Prepared by Rozendal J. A. (1997). Available from: https://www.who.int/whopes/resources/vector_rozendaal/en/ Acess: 04/09/2020.

42. StataCorp: Stata Statistical Software: Release 15. College Station, TX: StataCorp LLC. https://www.stata.com/ (2017).

43. Dias JC, Silveira AC, Schofield CJ. The impact of Chagas disease control in Latin America: a review. Mem Inst Oswaldo Cruz. 2002;975:603–12. doi: 10.1590/s0074-02762002000500002.

44. Moncayo A. Chagas disease: Current epidemiological trends after the interruption of vectorial and transfusional transmission in the southern cone countries. Memorias Do Instituto Oswaldo Cruz. 2003;98 5:577–91; doi: 10.1590/s0074-02762003000500001. WOS:000184766900001.

45. Espinoza N, Borras R, Abad-Franch F. Chagas disease vector control in a hyperendemic setting: the first 11 years of intervention in Cochabamba, Bolivia. PLoS Negl Trop Dis. 2014;8 4:e2782; doi: 10.1371/journal.pntd.0002782.

46. Fuseini G, Ismail HM, von Fricken ME, Weppelmann TA, Smith J, Logan RAE, et al. Improving the performance of spray operators through monitoring and evaluation of insecticide concentrations of pirimiphos-methyl during indoor residual spraying for malaria control on Bioko Island. Malaria Journal. 2020;19 1; doi: 10.1186/s12936-020-3118-y. WOS:000513868500005.

47. Huda MM, Mondal D, Kumar V, Das P, Sharma SN, Das ML, et al. Toolkit for monitoring and evaluation of indoor residual spraying for visceral leishmaniasis control in the Indian subcontinent: application and results. J Trop Med. 2011;2011:876742; doi: 10.1155/2011/876742.

48. Chowdhury R, Chowdhury V, Faria S, Islam S, Maheswary NP, Akhter S, et al. Indoor residual spraying for kala-azar vector control in Bangladesh: A continuing challenge. Plos Neglected Tropical Diseases. 2018;12 10; doi: 10.1371/journal.pntd.0006846. WOS:000449318100048.

49. Ismail HM, Kumar V, Singh RP, Williams C, Shivam P, Ghosh A, et al. Development of a Simple Dipstick Assay for Operational Monitoring of DDT. Plos Neglected Tropical Diseases. 2016;10 1; doi: 10.1371/journal.pntd.0004324. WOS:000372565700037.

50. Tan C-x, Shen D-l, Weng J-q, Chen Q-w, Liu H-j, Yuan Q-l. Research on the rheological properties of pesticide suspension concentrate. 2004. Available from: http://www.jzus.zju.edu.cn/article.php?doi=10.1631/jzus.2004.1604. Access: 04/09/2020.

51. Gorla D, Hashimoto K. Control strategies against Triatominae. In: Telleria J, Tibayrenc M, editors. American Trypanosomiasis Chagas Disease 884p, 2nd Edition edn: Elsevier Inc; 2017. p. 223–42.

52. Paz-Soldan VA, Bauer KM, Hunter GC, Castillo-Neyra R, Arriola VD, Rivera-Lanas D, et al. y To spray or not to spray? Understanding participation in an indoor residual spray campaign in Arequipa, Peru. Global Public Health. 2016;13 1:65–82; doi: 10.1080/17441692.2016.1178317. WOS:000419603300005.

53. Kumar V, Mandal R, Das S, Kesari S, Dinesh DS, Pandey K, et al. Kala-azar elimination in a highly-endemic district of Bihar, India: A success story. Plos Neglected Tropical Diseases 2020; 14 (5): e0008254 doi:10.1371/journal.pntd.0008254.

54. Thawer NG, Ngondi JM, Mugalura FE, Emmanuel I, Mwalimu CD, Morou E, et al. Use of insecticide quantification kits to investigate the quality of spraying and decay rate of bendiocarb on different wall surfaces in Kagera region, Tanzania. Parasites & Vectors. 2015;8; doi: 10.1186/s13071-015-0859-5. WOS:000354237100001.

55. Kaur H, Allan EL, Eggelte TA, Monti F. A Colorimetric Test for the Evaluation of the Insecticide Content of LLINs Used on Bioko Island, Equatorial Guine. 2020. Available from: https://assets.researchsquare.com/files/rs-64354/v1/e2d6eccc-919a-4a30-9293-5cd027fb9dad.pdf. Access: 28/09/2020.

56. Schofield CJ. Comparison of sampling techniques for domestic populations of Triatominae. Transactions of the Royal Society of Tropical Medicine and Hygiene. 1978;72 5:449–55; doi: 10.1016/0035-9203(78)90160-8. WOS:A1978FW41900002.

57. Abad-Franch F, Valenca-Barbosa C, Sarquis O, Lima MM. All that glisters is not gold: sampling-process uncertainty in disease-vector surveys with false-negative and false-positive detections. PLoS Negl Trop Dis. 2014;8 9:e3187; doi: 10.1371/journal.pntd.0003187.

58. Rabinovich JE, Gurtler RE, Leal JA, Feliciangeli D. Density estimates of the domestic vector of Chagas disease, Rhocnius prolixus Stal (Hemiptera, Reduviidae), in rural houses in Venezuela. Bulletin of the World Health Organization. 1995;73 3:347-57. WOS:A1995RH59300009.

59. Provecho YM, Gaspe MS, Fernandez MD, Gurtler RE. House Reinfestation With Triatoma infestans (Hemiptera: Reduviidae) After Community-Wide Spraying With Insecticides in the Argentine Chaco: A Multifactorial Process. Journal of Medical Entomology. 2017;54 3:646–57; doi: 10.1093/jme/tjw224. WOS:000403153400015.

60. Cecere MC, Rodriguez-Planes LI, Vazquez-Prokopec GM, Kitron U, Gurtler RE. Community-based surveillance and control of chagas disease vectors in remote rural areas of the Argentine Chaco: A five-year follow-up. Acta Tropica. 2019;191:108–15; doi: 10.1016/j.actatropica.2018.12.038. WOS:000460495200014.

61. Gurevitz JM, Sol Gaspe M, Enriquez GF, Provecho YM, Kitron U, Guertler RE. Intensified Surveillance and Insecticide-based Control of the Chagas Disease Vector Triatoma infestans in the Argentinean Chaco. Plos Neglected Tropical Diseases. 2013;7 4; doi: 10.1371/journal.pntd.0002158. WOS:000318153100019.

62. Mougabure-Cueto G, Picollo MI. Insecticide resistance in vector Chagas disease: Evolution, mechanisms and management. Acta Tropica. 2015;149:70–85; doi: 10.1016/j.actatropica.2015.05.014. WOS:000359957500012.

63. Roca-Acevedo G, Ines Picollo M, Santo-Orihuela P. Expression of Insecticide Resistance in Immature Life Stages of Triatoma infestans (Hemiptera: Reduviidae). Journal of Medical Entomology. 2013;50 4:816–8; doi: 10.1603/me12116. WOS:000321897300017.

64. Carla Cecere M, Vazquez-Prokopec GM, Ceballos LA, Boragno S, Zarate JE, Kitron U, et al. Improved Chemical Control of Chagas Disease Vectors in the Dry Chaco Region. Journal of Medical Entomology. 2013;50 2:394–403; doi: 10.1603/me12109. WOS:000315997300025.

