## Additional file 1 laboratory assays for "Indoor residual spraying practises against *Triatoma infestans* in the Bolivian Chaco: contributing factors to suboptimal insecticide delivery to treated households"

**Laboratory assays**

*Chemical colorimetric assay IQK™*

Insecticide concentrations were quantified by colorimetric assay based on the chemistry of a commercial Insecticide Quantification Kit for Pyrethroids -IQK™ (ASSURE Cyano Pyrethroid Insecticides IVCC – AVIMA- Integrated Malaria Vector Control Solutions) designed to quantify or semi-quantify cyano-pyrethroid concentrations following others [38].

*Insecticide concentrations delivered onto filter papers*

Assay protocols and data analysis followed previously published methods [49]. Briefly, for each filter paper, the active ingredient was extracted from 2 filter paper punches (1.3cm$^2$ each, total area =2.6cm$^2$). To extract the pyrethroid, 800 µl of 0.075% potassium hydroxide [KOH] in 90% ethanol (Reagent A) was added to the glass tube containing the 2 punches, followed by the addition of 800 µl of reagent B to induce the colorimetric reaction. Reagent B was previously prepared diluting 0.4% 2,3,5-triphenyltetrazolium chloride [TTC] in 95% ethanol, later mixing it with a solution of 0.04% 4-nitrobenzaldehyde [PNB] in 95% ethanol and maintained in a glass bottle covered with aluminium foil. The sample was then vortexed for 1 minute followed by an incubation step of 10 minutes. The reaction was neutralized by adding 400 µl of 0.5% acetic acid diluted in 100ml of distillate water (Reagent C). The final reaction (200 µl) was placed in a plaque assay well, and the optical density (OD) of the colorimetric reaction measured using a photometer calibrated at 480nm wavelength. Optimization steps required adapting the described kit protocol by reducing (i) the filter paper punch sample area from 4cm$^2$ to 2.6cm$^2$, and (ii) the time of incubation from 15 to 10 minutes.

*Standardised insecticide concentration units*

Sample insecticide OD values were standardised across plates by comparing values to the standard curve, conducted for each assay. For filter paper delivered dose, 18 serial dilutions of alpha-cypermethrin a.i. were generated, corresponding to the filter paper sample punch area (ug/2.6cm$^2$), ranging from 0 to 80mg/m$^2$, using a stock concentration of alpha-cypermethrin at 0.5mg/ml.

For spray tank solutions, 6 serial dilutions were generated: 0.1875; 0.375; 0.75; 1.25; 2.5; 5 mg/ml of alpha-cypermethrin. The standard curve was generated using alpha-cypermethrin stock concentration at 10mg/ml.

The standardised insecticide concentration units (A), were calculated by

$$A = \frac{OD\ sample - OD\ control}{y\ value}$$

where y is the test sample units obtained from comparison to the standard curve, and A units are $\mu$g/2.6cm$^2$ (filter papers) or $\mu$l/ml (spray tank solutions). For the control, values were obtained in each plate by completing the control well with the IQK™ reagents only.

*IQK™ validation by high performance liquid chromatography (HPLC)*

To validate the colorimetric assay results, a random selection of filter paper samples was also tested by HPLC (n=27 filter papers from three sampled houses). Briefly, two filter paper punches (1.3cm$^2$ each= 2.6cm$^2$ total area) from each filter paper were placed inside a glass tube, and 5 ml of solvent added prepared from 100mg of standard dicyclohexyl phthalate [DCP] diluted in

900ml ethanol to a final concentration of 100 μg/ml), and vortexed for 1 minute. 1ml of the solution was then transferred to a new glass tube and vapored to dryness under nitrogen at 60ºC and stored at 4ºC overnight. Four glass tubes containing 1ml of the extraction solution alone were prepared as controls (DCP). To resuspend and clean the insecticide content, 1ml of methanol was then added to the cold stored glass tube and vortexed for 1 minute. 1ml of this sample was transferred to an Eppendorf tube and centrifuged for 20 minutes at 13,000rpm. A total of 250μl of the supernatant was transferred to an HPLC vial for HPLC analysis. A standard curve was prepared from alpha-cypermethrin dilution series 0, 62.25, 125, 250 and 1000μg/ml. HPLC analysis was performed by injection of 10μl aliquots of samples into a reverse-phase Hypersil GOLD C18 column (175 Å, 250 x 4.6mm, 5μm, Thermo Scientific, UK) at 23-25ºC. A mobile phase of acetonitrile/water 80:20 was used at a flow rate of 1ml/min. Alpha-cypermethrin peaks were detected at 232nm with an Ultimate 3000 UV detector (Dionex, Camberley, UK). Data was analyzed by Dionex Chromeleon software. Final concentrations in milligrams per square meter were estimated from the following equation:

$$A = \left(\frac{B}{C}\right) x\ V\ x\ D$$

where:

A= Alphacypermethrin in μg/sample

B= Peak Area (mAU*min)

C= Slope value (obtained from standard curve)

V= Volume of extraction solution added to the samples

D= Internal standard correction factor (obtained by dividing the DCP peak area by the average DCP area).
